# Knowledge, attitudes, and practices among the general population during COVID-19 outbreak in Iran: A national cross-sectional survey

**DOI:** 10.1101/2020.06.10.20127258

**Authors:** Edris Kakemam, Djavad Ghoddoosi-Nejad, Zahra Chegini, Khalil Momeni, Hamid Salehinia, Soheil Hassanipour, Hosein Ameri, Morteza Arab-Zozani

## Abstract

**Background:** COVID-19, which emerged in December 2019, is the largest pandemic ever to occur. During the early phase, little was known about public awareness relating to Coronavirus disease. This study was designed to determine knowledge, attitudes and practices (KAP) among the Iranian public towards COVID-19.

**Methods:** A cross-sectional online survey was carried out in Iran from 2 March to 8 April 2020 using a self-administered questionnaire on 1,480 people. COVID-19-related KAP questions were adapted from other internationally validated questionnaires specific to infectious diseases.

**Results:** All participants were aware of COVID-19. When asked unprompted, 80% of respondents could correctly cite fever, difficulty breathing and cough as signs/symptoms of COVID-19. Most of our sample population knew that by staying at home and staying isolated (95.3%, 95 % CI: 94.2-96.3) as well as constant hand washing and using disinfectants (92.5%, 95 % CI: 91.1-93.8) could prevent COVID-19. However, there was also widespread misconceptions such as the belief that COVID-19 can be transmitted by wild animals (58%, 95 % CI: 55.5-60.5) and by air (48.3%, 95 % CI: 45.7-50.8). Unprompted, self-reported actions taken to avoid COVID-19 infection included hand washing with soap and water (95.4%, 95 % CI: 94.3-96.4), avoiding crowded places (93%, 95 % CI: 91.7-94.3), cleaning hands with other disinfectants (80.9 %, 95 % CI: 78.9-82.9), and covering mouths and noses when coughing and sneezing (76.1 %, 95 % CI: 73.9-78.2). The internet and social media (94.5%, 95 % CI: 93.3-95.6) were the main Coronavirus information sources. However, the most trusted information sources on Coronavirus were health and medical professionals (79.3%, 95 % CI: 77.2-81.3). The majority of participants (77.0%, 95 % CI: 74.8-79.1) wanted more information about Coronavirus to be available.

**Conclusion:** Our findings suggest that people’s knowledge and attitude towards COVID-19 at the time of its outbreak was of a high level. Therefore, health systems should use multiple ways, such as mass media, phone applications, electronic, print, and tele-education to increase KAP related to COVID-19.

## Introduction

Having emerged in the last months of 2019, COVID-19 spread beyond an unimaginable rate, imposing a heavy burden on worldwide healthcare systems, forcing almost all countries to apply quarantine rules which were unprecedented over the last century. [1, 2].

Because of the characteristics of viral diseases and their transmission methods, which affect a huge number of people, COVID-19 received urgent attention globally. Traits such as high transmission rates, long incubation period - 7 to 14 days – and global spread, infecting hundreds of thousands of people, meant the virus needed to be tackled with careful design and planning, especially as in some cases a phobia began to emerge about this new exotic virus [3].

Primary results and examination have shown that the most important symptoms of Covid-19 are: fever, dry coughs, and shortness of breath. Having all three together, patients are advised to refer to healthcare facilities immediately, so that prognosis is quick. However, because we lack an appropriate vaccine or cure for treating and managing these patients, the best way in our current situation is paying special attention to disease prevention and breaking the transmission chain of the virus [4, 5].

Experts believe that disease prevention, in this case, requires personal and public hygiene. This includes washing hands with soap for at least twenty seconds, reducing interactions with other people to the minimum, staying home for at least 2 weeks after authorities have declared a lockdown, quarantining people who have come into contact with anyone infected as well as quarantining contaminated and infected residents in those areas with high infection rates. Actions like these needs a social awareness from both the authorities and the population at large, in order to handle the situation quickly and safely [1, 6].

In order to protect the public in Iran, it’s vitally important we understand the knowledge, attitude and practice (KAP) of people so that the authorities know which areas of KAP to target or enhance [7]. Therefore, having these statements in mind, it seems necessary to design and implement research activities in this area. This study, however, aims to assess the KAP of society about COVID-19. The results and evidence extracted from such surveys immediately help public health planners and other health sector authorities fight this virus in the most effective way.

## Materials and methods

### Study design

This cross-sectional survey was conducted from February 25 to April 25, to assess the public’s knowledge, attitudes, and practices regarding the Coronavirus outbreak in Iran.

### Population and sample size

The estimation of the sample size was done by assuming a minimum prevalence of 15%, confidence level□=□95%, and d□(margin of error)□=□0.02. The calculated sample size of this study was 1233 participants, and with design effect = 1.2 reaching a sample size of nearly 1480 participants.

### Measures

The survey questionnaire included both prompted direct questions as well as unprompted open-ended questions that allowed for multiple responses. Coronavirus KAP domains in the questionnaire included sources of information, knowledge about Coronavirus, behavioral intentions, prevention practices and attitudes towards survivors. The indicators used to assess Coronavirus KAP were informed by lessons learnt from similar KAP studies on other communicable diseases, especially MERS-COV, HIV/AIDS and Ebola [8-11]. Before the final survey was completed, changes were made as required to enable better understanding of the questions by the participants, and the arrangement of the questions was looked into to ensure its efficiency.

### Data collection

Due to maintaining social distance, the survey was done online and in all provinces of the country. The questionnaire was design on Porsline (similar to SurveyMonkey) shared on social media such as Telegram, WhatsApp, LinkedIn, and Facebook. People who were of Iranian nationality, were aged 10 years or more, and agreed to participate in the survey were directed to complete the questionnaire via clicking the link (https://survey.porsline.ir/#/). The survey started with a sentence that “completing the questionnaire by the participants is considered as voluntary participation”. After confirmation that participants understood this, people were guided to complete the self-report questionnaire. On average, questionnaires took approximately 10–15 minutes to complete. Our study was approved by the ethics committee of Birjand University of Medical Sciences before starting the formal survey (ethical code: IR.BUMS.REC.1398.391).

### Data analysis

Once data were collected, all questionnaires were then entered into a customized Excel-based system. All data were subsequently imported into and analyzed in SPSS version 23.0 (SPSS Inc., Chicago, Illinois, USA). Descriptive statistics were then generated for national-level estimates (proportions) and their 95% CIs.

## Results

A total of 1480 people responded to the questionnaire, yielding a response rate of 70.2%. The mean age was 31.29 years. Among the respondents, 797 (57.2%) were female and 782 (53.1%) were married. Of all respondents, 41.9% were between the ages 20-31years. Also, above 80% of respondents had an academic education.

### Awareness and risk perception

The majority of respondents (84.5%) had heard of Coronavirus disease 2019 (COVID-19) prior to the interview. Overall, 84.5% of respondents were aware that it is possible to survive and recover from COVID-19. Approximately 60% of respondents (59.6%) perceived themselves to be at some risk of contracting COVID-19.

### Knowledge of Coronavirus cause, transmission, signs and symptoms

In an open-ended question, the most common perceived cause/origin of Coronavirus was ‘virus’ (94.4%); only 31.7% linked Coronavirus to a ‘bats, monkeys, and wild animals’. Very few respondents mentioned that Coronavirus is caused by ‘Bacteria’ (6.7%) ‘Evildoing/Sin’ (4.1%) or ‘Parasites’ (3.7%).

In response to an unprompted open-ended question, the most frequently cited modes of transmission were: shaking hands with an infected person (91.9%), kissing and hugging (90.1%) and being in contact with the saliva of an infected person (87.2%).

Overall, 80% of respondents could name, without prompting, three key signs/symptoms of Coronavirus: ‘difficulty breathing’ (93.3%), ‘fever’ (90.9) and ‘cough’ (83.2%) (Table 2).

**Table 1:** Sociodemographic characteristics of respondents, (n=1480)

| Characteristic | N | %<sup>+</sup> |
| --- | --- | --- |
| **Sex\*** |  |  |
| Male | 597 | 42.8 |
| Female | 797 | 57.2 |
| **Age†** |  |  |
| ≤20 | 150 | 10.3 |
| 21-30 | 612 | 41.9 |
| 31-40 | 461 | 31.6 |
| 41-50 | 166 | 11.4 |
| >50 | 70 | 4.8 |
| **Marital Status‡** |  |  |
| Single | 692 | 46.9 |
| Married | 782 | 53.1 |
| **Educational Level§** |  |  |
| Lower Diploma | 51 | 3.5 |
| Diploma | 200 | 13.6 |
| Associate Degree | 115 | 7.8 |
| Bachelors | 595 | 40.3 |
| Master | 395 | 26.8 |
| PhD | 120 | 8.0 |
| **Region\*\*** |  |  |
| South | 144 | 9.8 |
| North | 412 | 28.0 |
| West | 411 | 27.9 |
| East | 288 | 19.6 |
| Center | 217 | 14.7 |

\*Missing values for sex=86.
†Missing values for age=21.
‡Missing values for marital Status =6.
§ Missing values for education=4.
\*\* Missing values for region =8.
+Valid percent

**Table 2:** Novel Coronavirus-related awareness, risk perceptions and knowledge (n=1480)

| Indicator | % | 95% CI |
| --- | --- | --- |
| **A-Awareness and risk perception** |  |  |
| Heard of Novel Coronavirus | 84.5 | 82.6-86.34 |
| Expressed that Novel Coronavirus existed in Iran | 98.1 | 97.4-98.8 |
| One of your family or relatives or friends infected by Coronavirus | 84.5 | 82.6-86.3 |
| Aware of possibility to have Novel Coronavirus without showing signs/symptoms | 79.5 | 77.4-81.5 |
| Aware of possibility to survive and recover from Novel Coronavirus | 84.5 | 82.6-86.3 |
| Aware of Novel Coronavirus call center to report sick persons and deaths | 52.8 | 50.2-55.3 |
| Perceived some risk of contracting Novel Coronavirus | 59.6 | 57.1-62.1 |
| **B—Knowledge of Novel Coronavirus cause** |  |  |
| Virus | 94.4 | 93.2-95.5 |
| Bats/ cats/ other wild animals | 31.7 | 29.3-34.1 |
| Parasites | 3.7 | 2.7-4.6 |
| Bacteria | 6.7 | 5.4-7.9 |
| Evildoing / Sin | 4.1 | 3.1-5.1 |
| I don't know/ not sure | 5.5 | 4.3-6.6 |
| **C—Knowledge of Novel Coronavirus modes of transmission** |  |  |
| Air | 44.6 | 42.1-47.1 |
| Eating 'animal meat' | 31.9 | 29.5-34.2 |
| Eating infected fruits and vegetable | 24.2 | 22.1-26.3 |
| Saliva of an infected person | 87.2 | 85.5-88.9 |
| Urine and faeces of an infected person | 37.3 | 34.8-39.7 |
| Breast milk of an infected person | 18.1 | 16.1-20.1 |
| Shaking hands with an infected person | 91.9 | 90.5-93.3 |
| Kissing and hugging | 90.1 | 88.6-91.6 |
| Others | 27.2 | 24.9-29.4 |
| **D—Knowledge of Novel Coronavirus signs and symptoms** |  |  |
| Knew three key signs/symptoms of Novel Coronavirus (fever, difficulty breathing, cough) | 80 | 77.9-82.1 |
| Vomiting | 28.4 | 26.1-30.7 |
| Diarrhea | 47.5 | 44.9-50.1 |
| Fever | 97.6 | 96.8-98.3 |
| Difficulty breathing (shortness of breathing) | 97.7 | 96.9-98.4 |
| Severe headache | 57.8 | 55.2-60.3 |
| Muscle pain | 59.1 | 56.6-61.6 |
| Cough | 94.3 | 93.1-95.4 |
| Sore throat | 45.1 | 42.5-47.6 |
| Abdominal pain | 15.9 | 14.1-17.7 |
| Weakness | 7.6 | 6.2-8.9 |

### Knowledge of Coronavirus prevention and treatment

Knowledge of Coronavirus prevention and treatment was high among respondents in that nearly everyone knew that Coronavirus can be prevented by staying at home and reducing contact with people (95.3%) as well as constant hand washing and using disinfectants (92.5%).

Furthermore, 75.1% of respondents expressed that early treatment of Coronavirus could increase survival and reduce the chance of transmission within the household (Table 3).

**Table 3:**
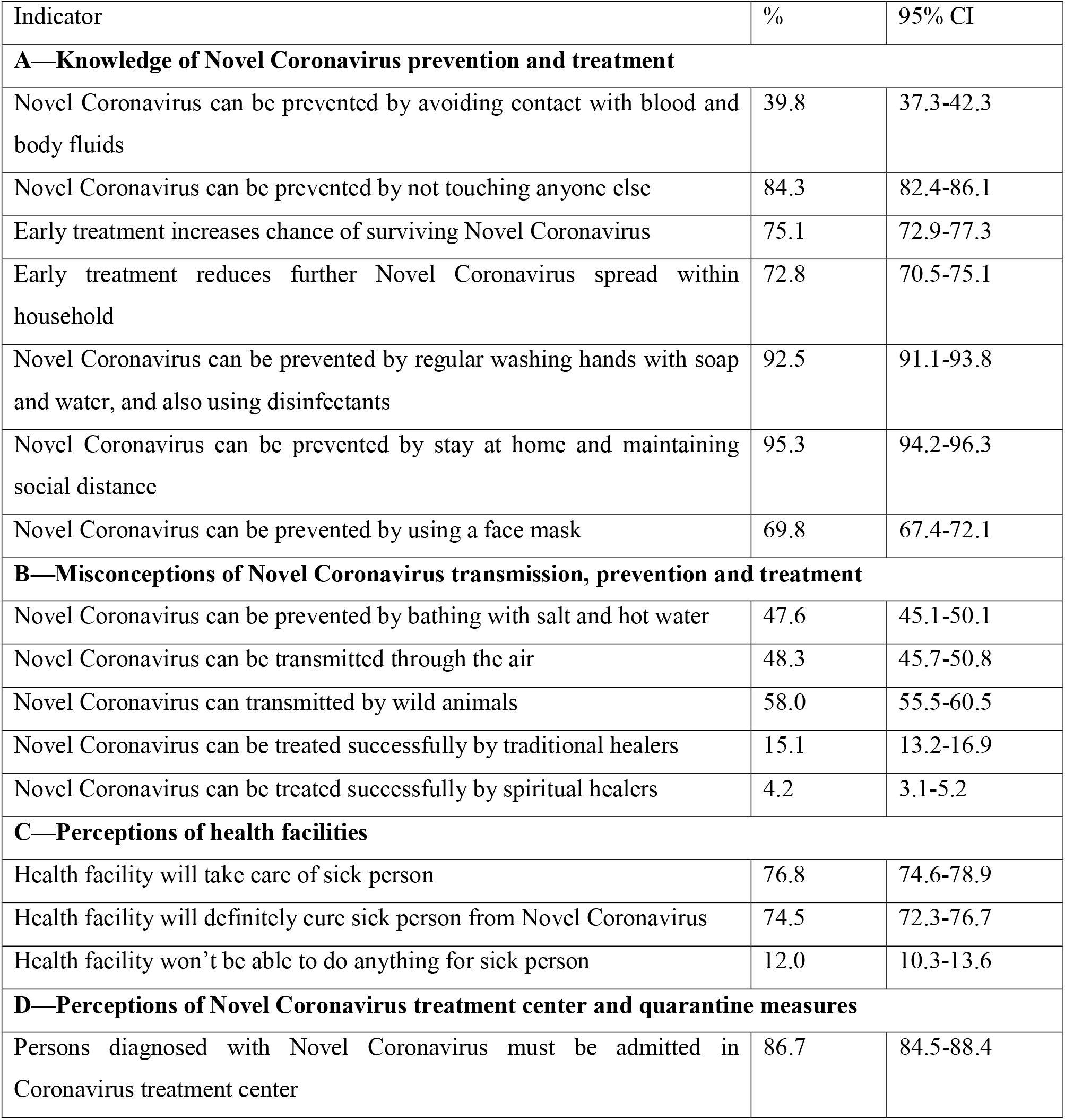
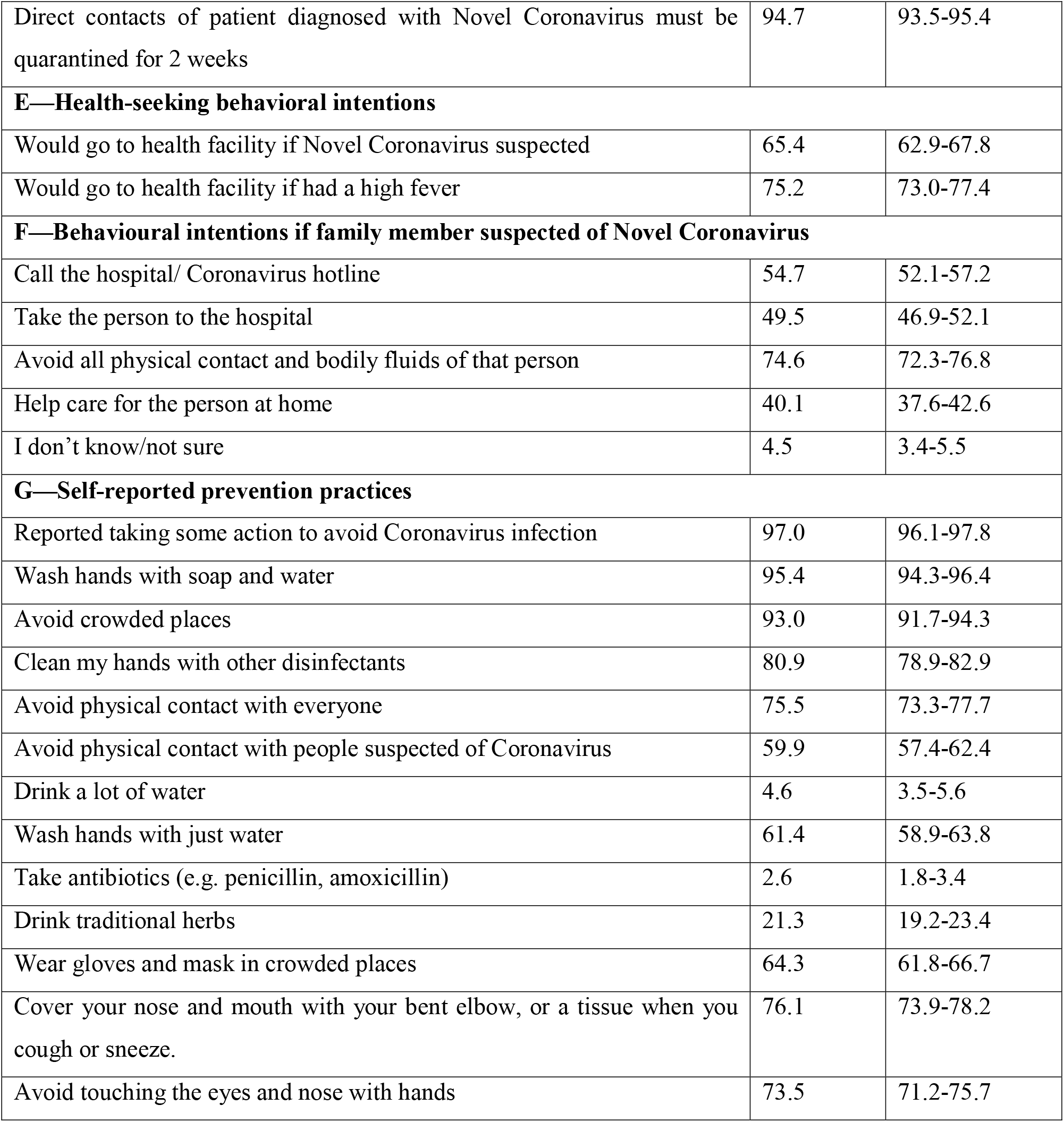
Novel Coronavirus - related knowledge, behavioral intentions and practices

| Indicator | % | 95% CI |
| --- | --- | --- |
| **A—Knowledge of Novel Coronavirus prevention and treatment** |  |  |
| Novel Coronavirus can be prevented by avoiding contact with blood and body fluids | 39.8 | 37.3-42.3 |
| Novel Coronavirus can be prevented by not touching anyone else | 84.3 | 82.4-86.1 |
| Early treatment increases chance of surviving Novel Coronavirus | 75.1 | 72.9-77.3 |
| Early treatment reduces further Novel Coronavirus spread within household | 72.8 | 70.5-75.1 |
| Novel Coronavirus can be prevented by regular washing hands with soap and water, and also using disinfectants | 92.5 | 91.1-93.8 |
| Novel Coronavirus can be prevented by stay at home and maintaining social distance | 95.3 | 94.2-96.3 |
| Novel Coronavirus can be prevented by using a face mask | 69.8 | 67.4-72.1 |
| **B—Misconceptions of Novel Coronavirus transmission, prevention and treatment** |  |  |
| Novel Coronavirus can be prevented by bathing with salt and hot water | 47.6 | 45.1-50.1 |
| Novel Coronavirus can be transmitted through the air | 48.3 | 45.7-50.8 |
| Novel Coronavirus can be transmitted by wild animals | 58.0 | 55.5-60.5 |
| Novel Coronavirus can be treated successfully by traditional healers | 15.1 | 13.2-16.9 |
| Novel Coronavirus can be treated successfully by spiritual healers | 4.2 | 3.1-5.2 |
| **C—Perceptions of health facilities** |  |  |
| Health facility will take care of sick person | 76.8 | 74.6-78.9 |
| Health facility will definitely cure sick person from Novel Coronavirus | 74.5 | 72.3-76.7 |
| Health facility won't be able to do anything for sick person | 12.0 | 10.3-13.6 |
| **D—Perceptions of Novel Coronavirus treatment center and quarantine measures** |  |  |
| Persons diagnosed with Novel Coronavirus must be admitted in Coronavirus treatment center | 86.7 | 84.5-88.4 |
| Direct contacts of patient diagnosed with Novel Coronavirus must be quarantined for 2 weeks | 94.7 | 93.5-95.4 |
| **E—Health-seeking behavioral intentions** |  |  |
| Would go to health facility if Novel Coronavirus suspected | 65.4 | 62.9-67.8 |
| Would go to health facility if had a high fever | 75.2 | 73.0-77.4 |
| **F—Behavioural intentions if family member suspected of Novel Coronavirus** |  |  |
| Call the hospital/ Coronavirus hotline | 54.7 | 52.1-57.2 |
| Take the person to the hospital | 49.5 | 46.9-52.1 |
| Avoid all physical contact and bodily fluids of that person | 74.6 | 72.3-76.8 |
| Help care for the person at home | 40.1 | 37.6-42.6 |
| I don't know/not sure | 4.5 | 3.4-5.5 |
| **G—Self-reported prevention practices** |  |  |
| Reported taking some action to avoid Coronavirus infection | 97.0 | 96.1-97.8 |
| Wash hands with soap and water | 95.4 | 94.3-96.4 |
| Avoid crowded places | 93.0 | 91.7-94.3 |
| Clean my hands with other disinfectants | 80.9 | 78.9-82.9 |
| Avoid physical contact with everyone | 75.5 | 73.3-77.7 |
| Avoid physical contact with people suspected of Coronavirus | 59.9 | 57.4-62.4 |
| Drink a lot of water | 4.6 | 3.5-5.6 |
| Wash hands with just water | 61.4 | 58.9-63.8 |
| Take antibiotics (e.g. penicillin, amoxicillin) | 2.6 | 1.8-3.4 |
| Drink traditional herbs | 21.3 | 19.2-23.4 |
| Wear gloves and mask in crowded places | 64.3 | 61.8-66.7 |
| Cover your nose and mouth with your bent elbow, or a tissue when you cough or sneeze. | 76.1 | 73.9-78.2 |
| Avoid touching the eyes and nose with hands | 73.5 | 71.2-75.7 |

### Misconceptions of Coronavirus transmission, prevention and treatment

There were widespread misconceptions about Coronavirus transmission, prevention and treatment. Nearly half (47%) of respondents said they could protect themselves from Coronavirus by washing with salt and hot water solution and nearly half again (48.3%) said Coronavirus is transmitted by air while 58% of the respondents expressed that Coronavirus is transmitted by wild animals. Moreover, 15.1% of respondents perceived that traditional healers could successfully treat Coronavirus (Table 3).

### Health-seeking behavioral intentions and self-reported prevention practices

Most of the respondents (75.2%) reported that they would go to a health facility if experiencing a high fever. Nearly all respondents (97%) reported taking some preventive action since learning about Coronavirus. Handwashing with soap and water was the most prevalent behavior reported in unprompted response (95.4%), followed by avoiding crowded places (93%), cleaning my hands with other disinfectants (80.9 %), and covering mouth and nose when coughing and sneezing (76.1 %).

### Attitudes towards Coronavirus survivors

Only 10.6% reported that they would not welcome back a Coronavirus survivor into the community though 49.5% would refuse to buy fresh vegetables from a shopkeeper who survived Coronavirus.

### Perceptions of Coronavirus vaccine and experimental treatment

73.2% of respondents reported they would accept an approved Coronavirus vaccine if it were to become available, while, only 26.1% said they would accept experimental treatments that have not been trialed (Table 4).

**Table 4:**
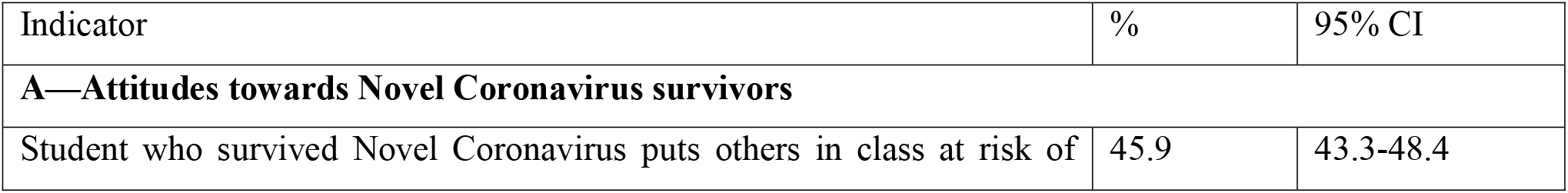
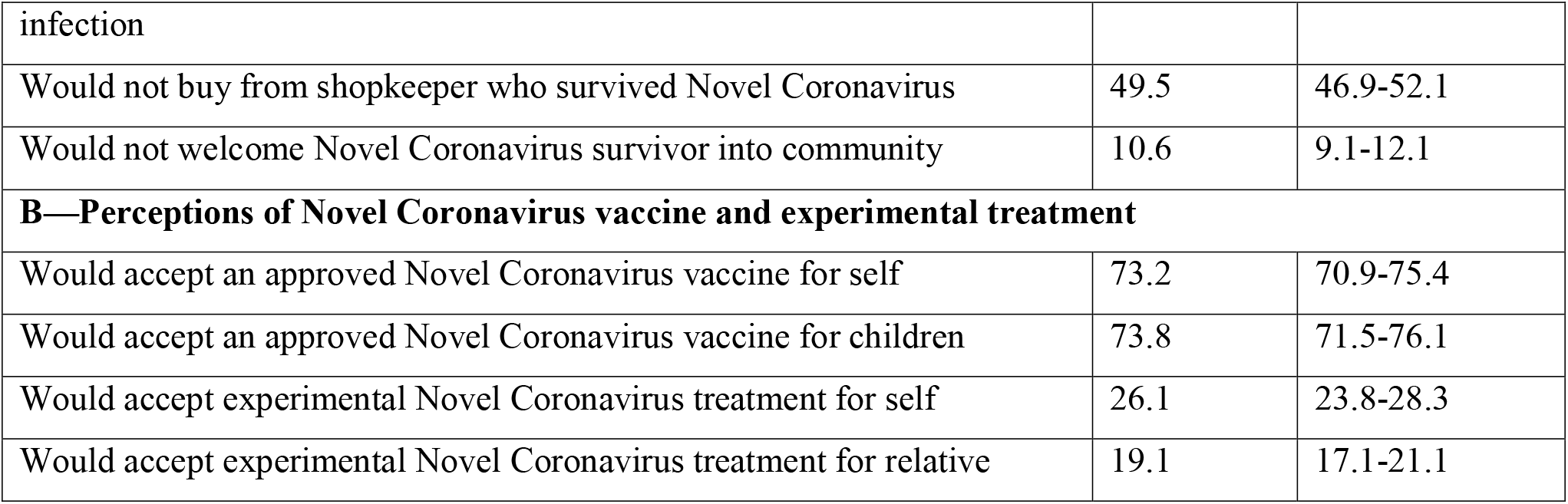
Attitudes towards Novel Coronavirus survivors and perceptions of Novel Coronavirus vaccine and experimental treatment

### Sources of receiving Coronavirus-related information

The internet and social media (94.5%) is the primary Coronavirus information channel mentioned by respondents in an open-ended question, followed by television (42%), relatives and friends (40.7%) and house visits by health workers (35.4%) (Table 5A). The most trusted information sources on Coronavirus were health and medical professionals (79.3%), the Government/Ministry of Health (55.7%) and the internet and social media (44.6%) (Table 5B).

**Table 5.** Novel Coronavirus -related information sources, trusted sources, and information gaps

| Indicator | % | 95% CI |
| --- | --- | --- |
| **A—Sources of receiving Novel Coronavirus -related information** |  |  |
| Internet and social media | 94.5 | 93.3-95.6 |
| Television | 70.7 | 68.3-73.0 |
| Relatives and friends | 40.7 | 38.2-43.2 |
| Radio | 14.6 | 12.8-16.4 |
| House visits by health workers | 35.3 | 32.8-37.7 |
| Community meetings | 4.3 | 3.2-5.3 |
| Mosque/other religious venues | 2.2 | 1.4-2.9 |
| Print materials | 10.4 | 8.8-11.9 |
| **B—Trusted sources of Novel Coronavirus -related information** |  |  |
| Health and medical professionals | 79.3 | 77.2-81.3 |
| Internet and social media | 44.6 | 42.1-47.1 |
| Government/Ministry of Health and Sanitation | 55.7 | 53.1-58.2 |
| Foreign media | 27.0 | 24.7-29.2 |
| Internal media | 33.9 | 31.5-36.3 |
| Relatives and friends | 7.1 | 5.8-8.4 |
| Traditional healers | 1.6 | 0.9-2.2 |
| Religious leaders | 1.8 | 1.1-2.4 |
| **C— Novel Coronavirus information gaps** |  |  |
| Wanted more Novel Coronavirus-related information on | 77.0 | 74.8-79.1 |
| Prevention | 62.2 | 59.7-64.6 |
| Medical care and treatment | 75.5 | 73.3-77.7 |
| Cause and origin | 47.2 | 44.6-49.7 |
| Signs and symptoms | 54.3 | 51.7-56.8 |

### Coronavirus information gaps

The majority of participants (77.0%) wanted more information about Coronavirus and specifically mentioned wanting to know more about providing medical care and treatment for infected people (75.5%) as well as how to prevent the disease (62.2%), while fifty-four respondents wanted more information on Coronavirus’ signs/symptoms and its cause/origin (47.2%) (Table 5C).

## Discussion

This study aimed to investigate the knowledge, attitudes and practices of the general population around COVID-19 in Iran and found that while the knowledge, attitudes and practices among the Iranian population are at a high level the acceptance of recovered Coronavirus patients in the community is rather low. In this regard, the KAP findings suggest that while during the outbreak, awareness and preventive behaviors related to COVID-19 were promoted well in Iran, misconceptions and discriminatory attitudes to those who survived Coronavirus were common. The study also highlighted the gap between perceived susceptibility of catching the virus and reality. For example, while almost all adults were at risk of catching Coronavirus, only 60% of people considered themselves at risk [12]. Therefore proper training about the susceptibility of all people and the possibility of getting the virus can increase awareness among individuals and help prevent infection. It is clear that each epidemic has its own unique characteristics; hence training in various areas of the disease, including susceptibility, is essential to prevent the spread of Coronavirus [13].

Knowledge about the signs and symptoms of Coronavirus disease was at a high level among people with over 80% providing correct answers. This compares with studies in various countries, including China[14], United States [15], and India [16], revealing that people are highly aware of Coronavirus, due to information in the mass media, including radio, television, social media and official authorities’ efforts like the Ministry of Health programs. Moreover, according to the results, 95% of people have stated that staying at home and obeying health protocols will prevent them from being infected. In this regard, studies have also illustrated that staying at home and maintaining social distance have a positive effect on reducing disease transmission [14]. Countries that did not enforce traffic regulations or imposed them later are suffering from a higher prevalence [17]. In order to maintain social distancing, several measures have been taken into account in Iran, including the prohibition of commuting on intercity routes, avoiding crowded places, and proroguing high-risk jobs. However, despite the numerous warnings of the government and the Ministry of Health, and also owing to the lack of effective laws, some people did not follow these measures.

In line with this, some misconceptions about the disease still persisted. For instance, 47% of people believe the virus is killed by saline solution, and 58% believe that the virus is transmitted through wild animals [18]. Any misconceptions about the disease and ways to prevent it could lead to a drastic increase in the incidence rate. Therefore, more detailed comprehensive training and information may need to be disseminated through the media, health practitioners, researchers and other stakeholders.

However, positive key findings show that 75% of people stated they would go to a health facility if they had a fever and 90% of people say they have taken preventive measures since the onset of Coronavirus such as washing their hands with soap and water and avoiding crowded areas.

One area of concern revealed by the study is the stigma of Coronavirus survivors. Some 10% of people said they would not welcome a survivor of Coronavirus into the community while 49.5% stated they would refuse to buy fresh vegetables from shopkeepers who had survived Coronavirus. This may raise main concerns about stigma attached to patients who have the disease among their community. Therefore, measures should be taken to increase awareness at the community level to reduce stigma [16, 19].

From the publicly available information sources, the most important source for acquiring knowledge for most people is social media. Therefore, it is necessary to provide accurate, precise and timely information to the public [20, 21].

Finally, more than 50% of people stated a need for more information and knowledge about treatment and diagnosis of the virus. Due to the fact that this is a new epidemic, it is important to have reliable and up-to-date information on all aspects of the disease; from prevention to diagnosis and treatment in order to acquire coherence and cohesion in people’s behavior using scientific protocols, not gossip or fake news in social media and on the internet.

### Strength and limitation

This study is the first to be conducted nationally in Iran about the KAPs of the general population regarding Coronavirus and has a high sample size. Since the study was conducted online and people who have access to virtual networks have been included in the study, it would be prudent to generalize the results to the whole community (people without smartphones and less literate).

## Conclusion

In conclusion, people’s knowledge, attitudes and performances about the signs and symptoms of the disease are at a high level, but there are misconceptions about the disease. Also, the fear of dealing with recovered patients and the lack of communication and knowledge around recovered patients are at a high level. However, the disease is unlikely to be transmitted to a person after recovery, so training and information via the media can be helpful in reducing misconceptions and the stigma around recovered patients in the community.

## Data Availability

The data is available if there is a reasonable request.

## List of abbreviations

COVID-19: Coronavirus disease 2019
KAP: Knowledge, Attitude and Practice
SARS-CoV-2: Severe acute respiratory syndrome coronavirus 2

## Declarations

### Ethics approval and consent to participate

This research was approved by Birjand University of Medical Sciences, Ethical Committee (Ethical code: IR.BUMS.REC.1398.391). During data collection, informed written consent was taken from participant.

### Consent for publication

Not applicable

## Availability of data and materials

### Funding

None

### Authors’ contributions

MA-Z and EK conceptualized the project. MA-Z, EK, DGh, HS and SH drafted the manuscript. EK and SH conducted data analysis. All authors reviewed and edited the manuscript. All authors read and approved the final version of the manuscript.

## Acknowledgements

We acknowledged Birjand University of Medical Sciences for supporting this research.

## References

1. Signorelli, C. and G. Fara, COVID-19: Hygiene and Public Health to the front. Acta Bio-medica: Atenei Parmensis, 2020. 91(3-S): p. 7–8.

2. Arab-Zozani, M. and S. Hassanipour, Features and Limitations of LitCovid Hub for Quick Access to Literature About COVID-19. Balkan medical journal, 2020.

3. Lipsitch, M., D.L. Swerdlow, and L. Finelli, Defining the Epidemiology of Covid-19—Studies Needed. New England Journal of Medicine, 2020.

4. Gostic, K., et al., Estimated effectiveness of symptom and risk screening to prevent the spread of COVID-19. eLife, 2020. 9: p. e55570.

5. Zu, Z.Y., et al., Coronavirus Disease 2019 (COVID-19): A Perspective from China. Radiology, 2020: p. 200490.

6. Abdelhafiz, A.S., et al., Knowledge, Perceptions, and Attitude of Egyptians Towards the Novel Coronavirus Disease (COVID-19). Journal of Community Health, 2020: p. 1–10.

7. Moftakhar, L. and M. Seif, The Exponentially Increasing Rate of Patients Infected with COVID-19 in Iran. Archives of Iranian medicine, 2020. 23(4): p. 235.

8. Jalloh, M.F., et al., National survey of Ebola-related knowledge, attitudes and practices before the outbreak peak in Sierra Leone: August 2014. BMJ global health, 2017. 2(4): p. e000285.

9. Shokoohi, M., et al., HIV knowledge, attitudes, and practices of young people in Iran: findings of a National Population-Based Survey in 2013. PLoS One, 2016. 11(9).

10. He, N., et al., Knowledge, attitudes, and practices of voluntary HIV counseling and testing among rural migrants in Shanghai, China. AIDS Education & Prevention, 2009. 21(6): p. 570–581.

11. Aldowyan, N., A.S. Abdallah, and R. El-Gharabawy, Knowledge, Attitude and Practice (KAP) Study about Middle East Respiratory Syndrome Coronavirus (MERS-CoV) among Population in Saudi Arabia. International Archives of Medicine, 2017. 10.

12. Wu, Z. and J.M. McGoogan, Characteristics of and important lessons from the coronavirus disease 2019 (COVID-19) outbreak in China: summary of a report of 72 314 cases from the Chinese Center for Disease Control and Prevention. Jama, 2020. 323(13): p. 1239–1242.

13. Johnson, E.J. and S. Hariharan, Public health awareness: knowledge, attitude and behaviour of the general public on health risks during the H1N1 influenza pandemic. Journal of Public Health, 2017. 25(3): p. 333–337.

14. Zhong, B.-L., et al., Knowledge, attitudes, and practices towards COVID-19 among Chinese residents during the rapid rise period of the COVID-19 outbreak: a quick online cross-sectional survey. International Journal of Biological Sciences, 2020. 16(10): p. 1745.

15. Clements, J.M., Knowledge and behaviors toward COVID-19 among US residents during the early days of the pandemic. medRxiv, 2020.

16. Roy, D., et al., Study of knowledge, attitude, anxiety & perceived mental healthcare need in Indian population during COVID-19 pandemic. Asian Journal of Psychiatry, 2020: p. 102083.

17. McCloskey, B., et al., Mass gathering events and reducing further global spread of COVID-19: a political and public health dilemma. The Lancet, 2020. 395(10230): p. 1096–1099.

18. Zhou, M., et al., Knowledge, attitude and practice regarding COVID-19 among health care workers in Henan, China. Journal of Hospital Infection, 2020.

19. Organization, W.H., Rolling updates on coronavirus disease (COVID-19).[cited 2020 April 14] Available at: https://www.who.int/emergencies/diseases/novel-coronavirus-2019/events-as-they-happen. 2020, Accessed.

20. Almutairi, K.M., et al., Awareness, attitudes, and practices related to coronavirus pandemic among public in Saudi Arabia. Family & community health, 2015. 38(4): p. 332–340.

21. Kharma, M.Y., et al., Assessment of the awareness level of dental students toward Middle East Respiratory Syndrome-coronavirus. Journal of International Society of Preventive & Community Dentistry, 2015. 5(3): p. 163.

